# DECREASED VARIATION IN ESOPHAGEAL ACID CONCENTRATION IN DIFFERENT PHENOTYPES OF SYMPTOMATIC GASTROESOPHAGEAL REFLUX DISEASE

**DOI:** 10.1101/2023.03.31.23288021

**Authors:** Jerry D. Gardner

**Author notes:** The work described in this manuscript has not been published and is not being considered for publication elsewhere. **Ethics Approval:** For this retrospective analysis of clinically indicated tests with no identifiable patient data, the Stanford University Institutional Review Board determined that this research does not involve human subjects as defined in 45 CFR 46.102(f) or 21 CFR 50.3 (g). All data produced in the present study are available upon reasonable request to the authors.

## Abstract

**Background:** Homeostatic physiologic systems are regulated to maintain a particular variable such as blood pressure within a restricted range with relatively low variation, and this variation is increased with disease. Fractal physiologic systems, on the other hand, are characterized by wide, complex variation of variables such as heart rate or the walking stride interval, and this variation is decreased with disease. The present report examines time-series recordings of esophageal pH from normal subjects and different GERD phenotypes to measure the distributions of esophageal pH values and the distributions of changes in esophageal acid concentrations.

**Methods:** Using Lyon consensus definitions of symptomatic GERD phenotypes, I analyzed 24-hour esophageal pH recordings from normal subjects (n=20), Functional Heartburn subjects (n=20), Reflux Hypersensitivity subjects (n=20), and nonerosive esophageal reflux disease (NERD) subjects (n=20). For each subject I calculated the distribution of pH values as well as the distribution of changes in esophageal acid concentrations.

**Results:** Esophageal pH values have a power law distribution in both normal and symptomatic GERD phenotypes, and esophageal acid concentrations vary over four orders of magnitude in each group. The variation in esophageal acid concentrations decreased progressively from normal subjects to Functional Heartburn subjects to Reflux Hypersensitivity subjects to NERD subjects.

**Conclusions:** The decreased variation in esophageal acid concentration in symptomatic GERD phenotypes represents changes associated with disease in a fractal physiologic system that shares characteristic features with other fractal systems where disease states have been associated with decreased variation of heart rate or walking stride interval.

## INTRODUCTION

Frequently, disease or ageing are accompanied by changes in one or more variables in physiologic systems (1-4). Homeostatic physiologic systems are regulated to maintain a particular variable such as blood pressure within a restricted range with relatively low variation, and this variation is increased with ageing or disease (1, 2). Fractal physiologic systems, on the other hand, are characterized by wide, complex variation of variables such as heart rate or the walking stride interval, and this variation is decreased with ageing or disease (3, 4).

The Lyon Consensus Conference (5, 6) proposed criteria for the clinical diagnosis of three different phenotypes of gastroesophageal reflux disease (GERD): nonerosive gastroesophageal reflux disease (NERD), Reflux Hypersensitivity, and Functional Heartburn. Previous analyses, before recognition of different phenotypes of GERD, found that time-series of esophageal pH showed a fractal pattern in normal and GERD subjects (7). In other analyses esophageal pH values had a power law distribution in normal and GERD subjects (8).

The present report examines time-series recordings of esophageal pH from normal subjects and different GERD phenotypes to measure the distributions of esophageal pH values as well as the distributions of changes in esophageal acid concentrations.

## SUBJECTS

Patients were identified by exploring the electronic database at the Royal London Hospital GI Physiology Unit that contains clinically indicated impedance-pH recordings (Sandhill Scientific, Highlands Ranch, CO) from patients with typical symptoms of gastroesophageal reflux.

Using Lyon consensus definitions of symptomatic GERD phenotypes,(5, 6), I selected 24-hour esophageal pH recordings from normal subjects (n=20), Functional Heartburn subjects (n=20), Reflux Hypersensitivity subjects (n=20), and nonerosive esophageal reflux disease (NERD) subjects (n=20). All subjects had a normal upper gastrointestinal endoscopy at the time of the impedance-pH study. The esophageal pH recordings from one normal subject were technically unsatisfactory and were omitted from the present analyses.

For this retrospective analysis of clinically indicated tests with no identifiable patient data, the Stanford University Institutional Review Board determined that this research does not involve human subjects as defined in 45 CFR 46.102(f) or 21 CFR 50.3 (g) (9).

Values from impedance-pH testing from subjects for the present analyses have been published previously (10-12). Normal subjects, Functional Heartburn subjects and Reflux Hypersensitivity subjects all had normal esophageal acid exposure time (AET) with esophageal pH <4 for less than 4% of the 24-hour esophageal pH recording. NERD subjects had increased esophageal AET of pH <4 for greater than 6% of the 24-hour esophageal pH recording. Reflux Hypersensitivity subjects had a positive association of symptoms with reflux episodes (13, 14), whereas Functional Heartburn subjects had no association of symptoms with reflux episodes. Normal subjects had no symptoms during the impedance-pH testing.

## METHODS

The impedance-pH catheter was inserted with an esophageal pH sensor positioned 5cm above the upper border of the lower esophageal sphincter. Six impedance channels were positioned 3, 5, 7, 9, 15 and 17cm above the upper border of the lower esophageal sphincter, respectively. Software provided by Sandhill to process pH recordings automatically adjusts all pH values for the difference between the calibration temperature of 25C and the recording temperature of 37C. Software provided by Sandhill was also used to export pH data for every 4^th^ second of the recording to an Excel file.

All pH values below 0.5 were replaced by 0.5 and all pH values above 7.5 were replaced by 7.5.

To determine cumulative acid concentration for a subject, all pH values were converted to acid concentration in mmol/L Cumulative acid concentration was calculated as the sum of all values for acid concentration for that subject, and each value of acid concentration was then expressed as a percentage of the cumulative acid concentration. The distribution of the change in sequential values of acid concentration was calculated for esophageal acidity for each subject. The frequency distributions of changes in acid concentration for each group of subjects were calculated as the means of the values from all subjects in the group for each bin of the distribution. Expressing acid concentration as a percentage of cumulative acid concentration for a given subject makes it possible to examine differences in the distributions of changes in acid concentration that do not depend on the magnitude of the acid concentration per se.

Others (1) have calculated the change in values in a time-series as a percentage difference from the mean of all values.

Curve fitting and statistical analyses were performed using GraphPad Prism 9.4.1 software. Because the present analyses were exploratory, P-values were not adjusted for multiple comparisons.

## RESULTS

Figure 1 shows a linear relationship between the values of esophageal pH and the logarithm of the frequency of the values for Normal subjects and each GERD phenotype. Results in Table 1 show that each slope of the lines in Figure 1 is significantly different from zero and accompanying values of R^2^ are at least 0.90. The linear relationships illustrated in Figure 1 are described by a power law in that the frequency of what is being measured is a negative exponential function of the magnitude of what is being measured, and the value of the exponential is given by the slope. The term “power law” is used because the frequency of a particular pH value is a power of the pH value.

**Figure 1.**
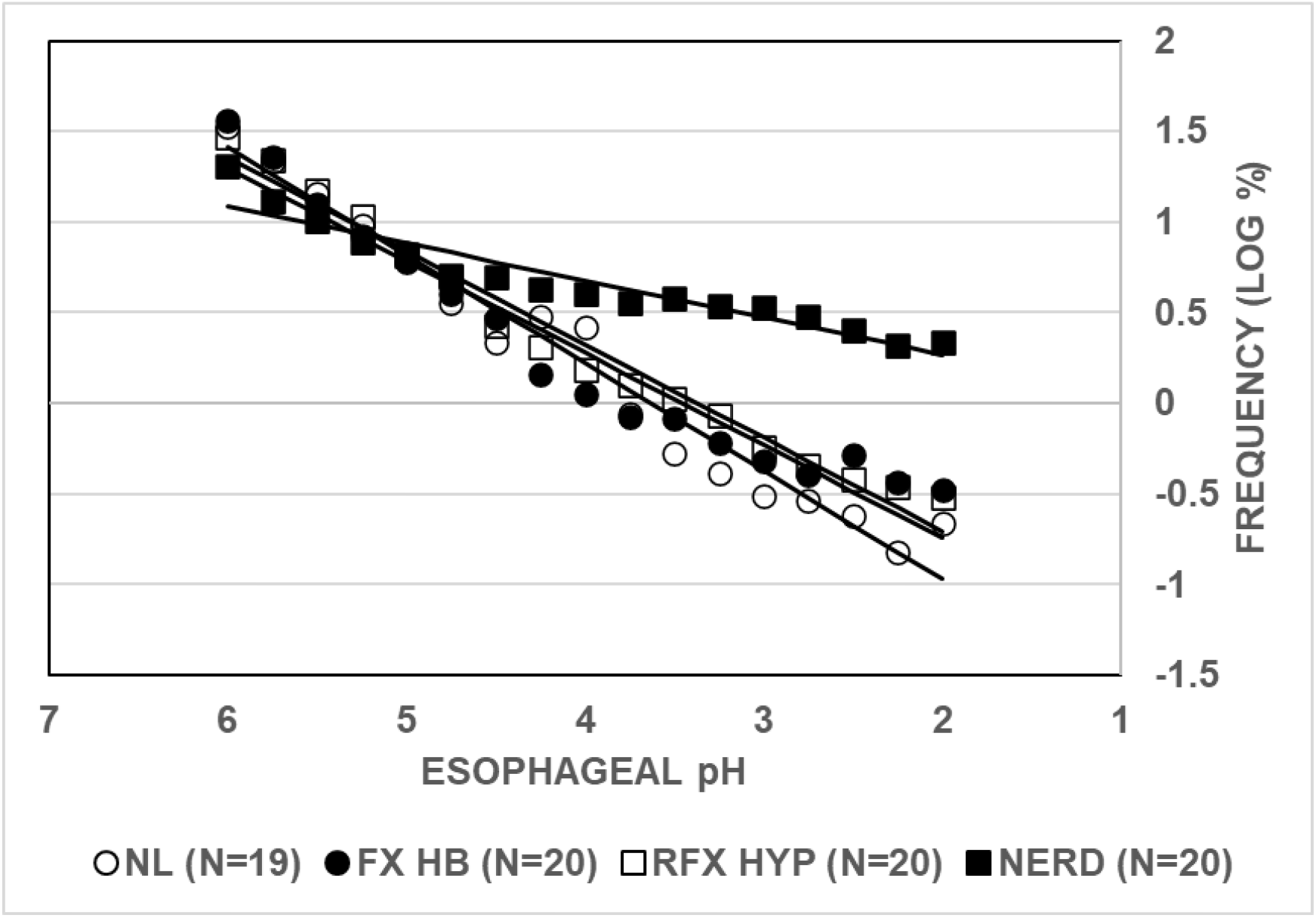
Distributions of values of esophageal pH in normal subjects and GERD phenotypes. Values given on the Y-axis are mean frequencies for the bin indicated on the X-axis. The solid lines are from linear, least-squares regression analyses. Abbreviations are NL – normal subjects; FX HB – Functional Heartburn subjects; RFX HYP – Reflux Hypersensitivity subjects; NERD – Nonerosive Reflux Disease subjects. The number of subjects in each group is given in parentheses after each abbreviation.

**Table 1.** RESULTS FROM LINEAR REGRESSION ANALYSES OF THE DISTRIBUTIONS OF ESOPHAGEAL pH VALUES.

|  | NORMAL | FUNCTIONAL<br>HEARTBURN | REFLUX<br>HYPERSENSITIVITY | NERD |
| --- | --- | --- | --- | --- |
| SLOPE | -0.5958 | -0.5094 | -0.5154 | -0.2047 |
| R<sup>2</sup> | 0.966 | 0.936 | 0.976 | 0.900 |
| P-VALUE<br>SLOPE NON-ZERO | <0.0001 | <0.0001 | <0.0001 | <0.0001 |
| P-VALUE SLOPE NON-ZERO was determined with an F-test. Results are from linear, least-squares regression analyses of the data in Figure 1 |  |  |  |  |

Table 2 shows results from pairwise comparisons of the values of the slopes given in Table 1. The slope from normal subjects was significantly higher than that from Reflux Hypersensitivity subjects and NERD subjects. It seemed possible that the lack of statistical significance comparing the slope from Functional Heartburn subjects to that from normal subjects might be due to an under-powered sample size. The slope from NERD subjects was significantly lower than that from normal subjects, from Functional Heartburn subjects, and from Reflux Hypersensitivity subjects. The slope from Functional Heartburn subjects was not significantly different from that from Reflux Hypersensitivity subjects.

**Table 2.** COMPARISON OF THE SLOPES OF DISTRIBUTIONS OF ESOPHAGEAL pH VALUES IN NORMAL SUBJECTS AND GERD PHENOTYPES.

| COMPARISON | P-VALUE | F-VALUE (DFn, DFd) |
| --- | --- | --- |
| ARE SLOPES DIFFERENT? | <0.0001 | 43.29 (3, 60) |
| NL VS FX HB | 0.0633 | 3.72 (1, 30) |
| NL VS RFX HYP | 0.0305 | 5.16 (1, 30) |
| NL VS NERD | <0.0001 | 135.0 (1, 30) |
| FX HB VS RFX HYP | 0.8823 | 0.022 (1, 30) |
| FX HB VS NERD | <0.0001 | 62.24 (1, 30) |
| RFX HYP VS NERD | <0.0001 | 130.7 (1, 30) |
| Abbreviations are NL – normal; FX HB – functional heartburn; RFX HYP – reflux hypersensitivity; NERD – non erosive reflux disease; DFn – degrees of freedom numerator; DFd – degrees of freedom denominator. The p-value is from an F-test. |  |  |

Figure 2 shows that the distribution of changes in cumulative esophageal acidity is biphasic for each GERD phenotype and normal subjects, and each distribution spans at least 4 orders of magnitude. The distribution of changes was significantly better fit by a 6^th^ order polynomial than by a 5^th^ order polynomial (P<0.0001 by F-test). A 7^th^ order polynomial did not give a significantly better fit than a 6^th^ order polynomial by an F-test. The polynomial model has no physiologic significance. It is simply a model that creates a curve that comes close to the data points and makes it possible to test the data for statistical differences.

**Figure 2.**
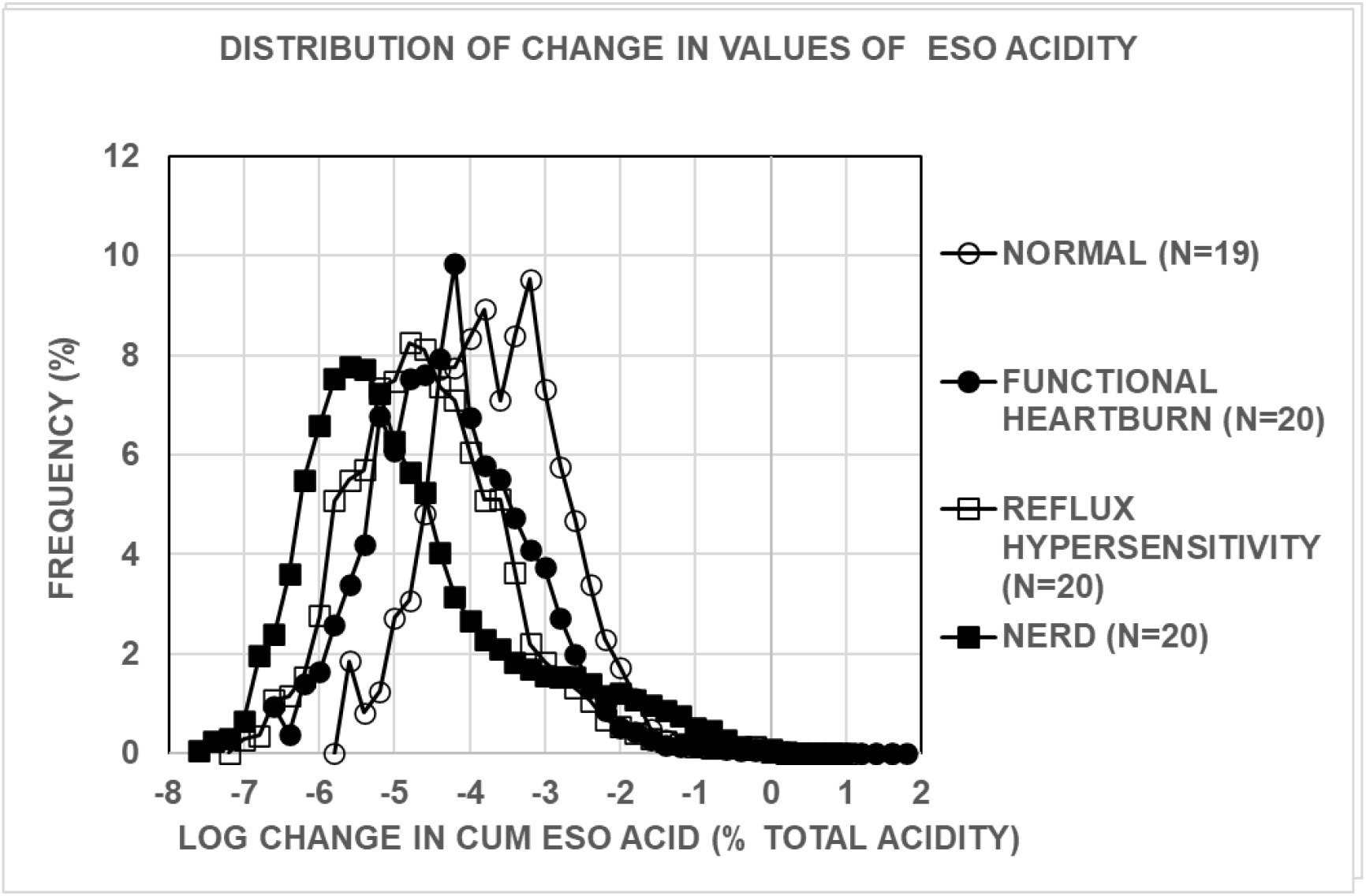
Distributions of change in values for cumulative esophageal acidity for GERD phenotypes and normal subjects. Values given on the Y-axis are mean frequencies for the bin indicated on the X-axis. Abbreviations are CUM – cumulative; ESO – esophageal.

Table 3 gives results from statistical comparisons of the data for distributions of changes in esophageal acid concentration in Figure 2 and indicates that all pairwise comparisons are significantly different at P<0.0001 by an F-test. Furthermore, the variation in esophageal acid concentrations is indicated by the range of values for change in esophageal acid concentration and is greatest in normal subjects, less in Functional Heartburn, still less in Reflux Hypersensitivity and least in NERD.

**Table 3.** COMPARISON OF THE DISTRIBUTIONS OF CHANGES IN ESOPHAGEAL ACIDITY IN GERD PHENOTYPES USING FITS TO A 6^TH^ ORDER POLYNOMIAL.

|  |  |  |
| --- | --- | --- |
| **COMPARISON** | **P-VALUE** | **F-VALUE (DFn, DFd)** |
| DIFFERENT CURVE FOR AT LEAST 1 DATASET | <0.0001 | 49.03 (21, 138) |
| NL VS FX HB | <0.0001 | 33.25 (7, 67) |
| NL VS RFX HYP | <0.0001 | 69.32 (7, 66) |
| NL VS NERD | <0.0001 | 90.87 (7, 67) |
| FX HB VS RFX HYP | <0.0001 | 9.827 (7, 71) |
| FX HB VS NERD | <0.0001 | 47.94 (7, 72) |
| RFX HYP VS NERD | <0.0001 | 29.01 (7, 71) |
| Abbreviations are NL – normal; FX HB – functional heartburn; RFX HYP – reflux hypersensitivity; NERD – non erosive reflux disease; DFn – degrees of freedom numerator; DFd – degrees of freedom denominator. The p-value is from an F-test. |  |  |

## DISCUSSION

The present analyses of time-series of esophageal pH measurements show that esophageal pH values have a power law distribution in both normal and symptomatic GERD phenotypes, and that esophageal acid concentrations vary over four orders of magnitude in each group. Also, the variation in esophageal acid concentration decreased progressively from normal subjects to Functional Heartburn subjects to Reflux Hypersensitivity subjects to NERD subjects. Thus, the different GERD phenotypes based on clinical characteristics identified by the Lyon Consensus Conference (5, 6) may actually share a common pathophysiology of varying severity. The decreased variation in esophageal acid concentrations in symptomatic GERD phenotypes represents changes associated with disease in a fractal physiologic system that is characterized by wide, complex variation under normal circumstances. Thus, GERD phenotypes share characteristic features with other fractal systems where disease states have been associated with decreased variation of heart rate or walking stride interval (3, 4).

All GERD phenotypes have symptoms of gastroesophageal reflux such as heartburn, regurgitation, or chest pain (5, 6) even though Functional Heartburn subjects and Reflux Hypersensitivity subjects have normal esophageal acid exposure times. It seems possible that regardless of the value of esophageal acid exposure time, the decreased variation in esophageal acid concentration, which can result in runs of self-similar esophageal acid concentrations, may represent a pathophysiologic signal to esophageal mucosa that triggers a symptom. Previous analyses of the same subjects used for the present study found that esophageal acid sensitivity appears to oscillate in each GERD phenotype, and for a given value of esophageal acid sensitivity, Reflux Hypersensitivity subjects have significantly more sequential symptoms associated with this sensitivity than do Functional Heartburn subjects (11). This difference between Functional Heartburn subjects and Reflux Hypersensitivity subjects might indicate that the lower variation of values of esophageal acid concentrations in Reflux Hypersensitivity subjects can cause an increase in GERD symptoms.

## Data Availability

All data produced in the present study are available upon reasonable request to the authors

## Acknowledgement

I am grateful to Dr. Daniel Sifrim, Director of Upper GI Physiology Unit, Royal London Hospital for providing the impedance-pH records and for stimulating, helpful discussions.

